# Potential Cost of Multi-Cancer Detection Tests to Medicare

**DOI:** 10.64898/2026.08.24.26361257

**Authors:** Laura D. Scherer, Daniel D. Matlock, John Cronin, Mark Gritz

## Abstract

Multi-Cancer Detection (MCD) tests can detect more than 50 different types of cancer using a blood test. Recently passed law in the U.S. guarantees that Medicare will pay for these tests when they are FDA approved and show evidence for clinical benefit. This manuscript provides estimates of the cost of MCD tests to Medicare under different assumptions of cost per test, eligibility, and screening uptake in the eligible population. This manuscript additionally estimates the cost of follow-up testing resulting from false positive results, which are considered avoidable costs caused by the screening test.

## Introduction

Multi-Cancer Detection (MCD) tests can detect more than 50 different types of cancer using a blood test.^1^ Trials have shown that MCDs have potential for early cancer detection, although they are more sensitive for advanced cancers (stage III/IV) compared to early-stage cancers (stage I/II).^2^ To date, no trial has shown evidence that MCDs offer a mortality benefit or favorable shift in cancer stage. No MCD test has received FDA approval, however, some tests can be legally marketed and ordered.

In 2026 the Nancy Gardner Sewell Medicare Multi-Cancer Early Detection Screening Coverage Act (H.R. 842) was signed into law. This law allows Medicare coverage and payment for annual MCD tests for beneficiaries 67 years and younger, provided that the MCD test is approved by the Food and Drug Administration (FDA) and shown to have clinical benefit. The FDA is poised to vote on premarket approval for the MCD Galleri^TM^ in September 2026. The purpose of this report is to estimate the potential cost of these tests to Medicare.

A second goal of this report is to estimate the potential cost of follow-up tests triggered by false positive MCD tests. Patients with a “cancer signal detected” MCD result require additional diagnostic testing to determine whether cancer is present and, if so, to identify its location.^2^ This distinguishes MCD screening from organ-specific screening for cancers such as breast, colorectal, and lung cancer, where an abnormal screening result already points to the organ requiring further evaluation. With true positive results, these additional diagnostic tests may lead to a benefit (e.g., cancer death prevented; although there is currently no evidence that MCD tests offer mortality benefit). By contrast, false positive results offer no health benefit, and additional tests that are triggered by a false positive result can be characterized as avoidable healthcare costs caused by the MCD test.

## Methods

### Estimating the annual cost of MCD testing to Medicare

The cost of MCD tests to Medicare was assumed to be a “preventive service;” therefore all costs would be borne entirely by Medicare and there are no out-of-pocket (OOP) payments for these services required from beneficiaries. Cost of MCD testing was estimated using two prices: (1) current price of Grail’s Galleri^TM^ test ($949), and (2) the average 2026 payment amount for a multi-target stool screening DNA test ($505) that is the benchmark in the Nancy Gardner Sewell Act.

Currently it is unclear who would be eligible for testing, therefore we provide cost estimates for two eligibility age ranges. The law specifies payment for annual screening for beneficiaries under the age of 68 as of January 2028 and increases the age limit by one year every calendar year thereafter. To estimate the approximate number of Medicare beneficiaries that would be eligible for testing, we used the 2025 calendar year information from the Medicare Monthly Enrollment database and estimated the number of beneficiaries age 45 to 67 to be 15,907,774 using a simple proportional allocation of ages 65-67 from the 17,430,641 beneficiaries that are age 65 to 69. The age ranges included in MCD trials have typically been ages 50 and older. Therefore, we also estimated costs to Medicare if screening were expanded to ages 45 to 79, estimated to be 52,744,541 in 2025 (the upper threshold of 79 was selected because cancer screenings are often not recommended for people at advanced age).^3^ Although this population estimate includes some people who are younger than age 50, the under-50 group is small so that the total number is nonetheless a close estimate of the total relevant population of Medicare beneficiaries.

The cost of annual testing was estimated given these different prices and age ranges using two assumptions about MCED testing uptake among eligible Medicare beneficiaries. Analyses present scenarios in which testing uptake is similar to colorectal cancer screening (69% uptake among those eligible^4^), and to breast cancer screening uptake (78% of those eligible^5^).

### Estimating the cost to Medicare for follow-up testing triggered by false positives MCD tests

In the PATHFINDER trial, a total of 90 participants had a cancer signal detected and 57 of these were false positive results.^2^ The trial Appendix reports the different kinds of diagnostic tests that these 57 participants received. We identified procedure codes that corresponded to the types of procedures documented in the Appendix using clinical expertise. When the exact code was not obvious, we used a weighted average across multiple codes that correspond to the general procedure description in the Appendix.

Procedure cost estimates are based on the national average payment amounts reported in the Medicare Fee-For-Service Provider Utilization & Payment Data Physician & Other Practitioners Dataset public use file based on 2023 claims with separate amounts reported for office and facility places of service.^6^ The cost of each procedure code used in the analysis represents the national average allowed amount for services delivered in both an office setting and a facility setting. For the procedures delivered in a facility, we added in an estimate of the additional facility fees using the Medicare.gov Procedure Price Lookup tool^7^ for procedures with available information. The estimated cost of each procedure code was calculated using a weighted average of the cost for services delivered in office and facility settings where the weights were equal to the number of procedures delivered in each setting as reported in the Provider Utilization & Payment public use file. For procedures listed in the PATHFINDER Appendix that corresponded to multiple procedure codes (e.g. CT scan), the estimated cost was also calculated as a weighted average across all of the relevant procedure codes using the total number of services for each procedure code as the weight for that code. All cost measures were converted to 2025 dollars using the seasonally adjusted Personal Consumption Expenditures: Services: Health care chain-type price index.^8^

To apply these costs to the Medicare population we must make assumptions about the expected prevalence of false positive tests. For these population analyses we used the false positive rate from PATHFINDER 2, which reports a signal detection rate of 0.93% and a false positive rate of 38.4%.^9^ This false positive rate is substantially lower than the PATHFINDER 1 observed rate (63.3% of positive tests), thus these analyses provide an optimistic cost estimate.

## Results

**Table 1** shows that the total cost of the test alone (not including the cost of follow-up testing) is estimated at between $5,543 million and $39,043 million per year depending on MCED test price, eligibility age range, and uptake assumptions. To place these numbers into context, the annual estimated cost of all cancer screening services for the U.S population is estimated at $43 billion (not including the cost of follow-up testing), and $4 billion among Medicare-covered populations (in 2025 dollars).^10^ In short, providing MCD tests to Medicare beneficiaries would be more than the cost of all other cancer screening tests for these beneficiaries to almost 10 times the cost depending on the price of the MCD test, the number of Medicare beneficiaries eligible for the test and the uptake of MCD tests.

**Table 1.** Estimated cost of annual MCD testing in the Medicare population with different age eligibility criteria and uptake assumptions, in 2025 dollars.

| Eligibility age range | MCD Test Price | Assuming 69% uptake (colorectal cancer screening benchmark) | Assuming 78% uptake (breast cancer screening benchmark) |
| --- | --- | --- | --- |
|  |  | Cost of test only |  |
| Age 45-67 years (Nancy Gardner Sewell Act inclusion) | \$505 | \$5,543 million | \$6,266 million |
| | \$949 | \$10,417 million | \$11,775 million |
| Age 45-79 years (typical MCD trial inclusion criteria) | \$505 | \$18,379 million | \$20,776 million |
| | \$949 | \$34,538 million | \$39,043 million |
|  |  | **Cost of follow-up tests triggered by false positives\*** |  |
| Age 45-67 years (Nancy Gardner Sewell Act inclusion) | | \$56 million Medicare<br>\$14 million Out of Pocket | \$64 million Medicare<br>\$16 million Out of Pocket |
| Age 45-79 years (typical MCD trial inclusion criteria) | | \$187 million Medicare<br>\$47 million Out of Pocket | \$212 million Medicare<br>\$53 million Out of Pocket |

\* Prevalence and type of follow-up tests were derived from PATHFINDER 1. Prevalence of false positive tests was derived from PATHFINDER 2, see main text.

For costs associated with follow-up testing after a false positive, we estimated that the average allowed amount (cost) per patient for follow-up testing after a false positive result in the PATHFINDER trial was approximately $1,799.69 (2025 dollars). Traditional Medicare beneficiaries without any supplemental insurance are responsible for a 20% co-insurance payment resulting in an average OOP cost of $359.94 (2025 dollars).

**Table 1** shows that the estimated total cost of follow-up tests triggered by false positive MCD tests is between $70 million and $265 million annually, depending on assumptions about eligibility and uptake. Traditional Medicare would pay 80% of these total costs and 20% would be paid out of pocket by beneficiaries. These are costs that would not have been incurred were it not for the initial MCD test and offer no health benefit to the patient (and may cause psychological distress and physical harm). Therefore, these costs can be considered excess costs of MCD testing.

## Discussion

Under the assumptions used in these analyses, the annual cost of MDC testing is estimated to be 1.4 to 9.8 times the current annual Medicare expenditures on all other preventive cancer screenings. Under assumptions of current out of pocket cost ($949), high uptake (78%) and broad eligibility (ages 45-79), the estimated cost of MCD testing for Medicare beneficiaries ($39 billion) approaches the total cost of all cancer screenings in the US population ($43 billion). For Medicare to pay for MCD tests and remain budget neutral under the assumptions of 69% uptake among individuals 79 and younger, MCDs would need to replace all other cancer screenings and the cost per test reduced to approximately $110.

It is reasonable to question the assumptions made in these analyses about the cost per test, eligibility, screening frequency, and uptake. MCD uptake may be lower in reality. Market competition may reduce the cost per test or Medicare could engage in price negotiation. The total cost could also be reduced by restricting eligibility or screening frequency (e.g., from annual to biennial). The estimated annual costs associated with follow-up testing from false positive tests is substantial ($70 million to $265 million), but small relative to the estimated cost of the MCD test itself.

A large investment in MCDs could come with the benefit of earlier detection for cancers that currently have no available screening test. However, MCD testing is unlikely to replace existing cancer screenings because MCD sensitivity for early-stage cancers is relatively poor. There is currently no evidence that MCD tests provide a reduction in cancer mortality or favorable shift in the prevalence of advanced cancers. Whether this significant financial investment will result in a population health benefit remains unknown.

## Data Availability

All data produced in the present work are contained in the manuscript

## Notes

### Competing Interest Statement

The authors have declared no competing interest.

### Funding

- Arnold Ventures: 25-13249

